# Predictive Value of Pulmonary Valve Annulus Z-Score for Valve Sparing in Tetralogy of Fallot Repair: A Systematic Review and Meta-Analysis

**DOI:** 10.1101/2025.01.31.25321449

**Authors:** Dicky Fakhri, Johan S. Sitanggang, Salomo Purba, Pribadi W. Busro, Budi Rahmat, Panji Utomo, Latifa Hernisa, Marshal B. Djaka, Doddy P. Pohan, Noverita S. Vinolina

## Abstract

**Background/Objectives:** Tetralogy of Fallot (ToF) is the most prevalent cyanotic congenital heart defect, requiring timely surgical intervention to improve survival. Two primary techniques for right ventricular outflow tract (RVOT) reconstruction are transannular patch (TAP) repair and valve-sparing (VS) surgery as a less invasive alternative. While TAP resolves pulmonary stenosis, it often results in long-term pulmonary regurgitation (PR). This meta-analysis investigates the pulmonary valve annulus (PVA) z-score as a predictor for choosing VS techniques to minimize complications, mean difference and cut-off analysis.

**Methods:** A systematic review and meta-analysis were conducted across 29 studies (N=5,806) assessing preoperative PVA z-scores in patients undergoing TAP or VS RVOT reconstruction. Data extraction followed PRISMA guidelines, with statistical analyses using a random-effects model by Review Manager 5.4.1 and receiver operating characteristic (ROC) curve evaluation.

**Results:** The meta-analysis showed significantly higher preoperative PVA z-scores in the VS group (MD: 1.01, 95% CI: 0.75–1.28, p < 0.001). The pooled grand mean PVA z-scores were −1.39 for VS and −2.97 for TAP. ROC analysis identified a z-score threshold of ≥ −2.59 (sensitivity: 88%, specificity: 80%) as optimal for maximizing VS surgery.

**Conclusions:** This study establishes the PVA z-score as a potential predictor for VS approaches in ToF repair, with a proposed threshold of ≥ −2.59 for optimal outcomes. Adoption of VS techniques guided by standardized z-score criteria may reduce PR-associated complications and enhance long-term survival and quality of life.

## 1. Introduction

Tetralogy of Fallot (ToF), the most prevalent cyanotic congenital heart disease (CHD), occurs in approximately 3.4 per 10,000 live births, accounting for 5% to 7% of all congenital heart anomalies. [1–3] Without prompt surgical intervention, the prognosis is poor, with only 10% of affected individuals surviving beyond 20 years of age, while the majority succumb to complications such as secondary myocardial hypertrophy and heart failure due to prolonged hypoxia. The preferred treatment strategy involves primary repair within the first year of life, with definitive corrective surgery being the optimal approach. The hallmark features of ToF include right ventricular outflow tract (RVOT) obstruction, ventricular septal defect, overriding of the aorta, and right ventricular hypertrophy. [4,5]

Complete surgical repair of Tetralogy of Fallot (ToF) has become standard practice during infancy, achieving an impressive post-repair survival rate of 95%-98% into adulthood. However, the timing of intervention and choice of surgical techniques can have profound and enduring effects on the need for future reinterventions, overall quality of life, and long-term survival of ToF patients.2 Reconstruction of the RVOT can be performed using either transannular patch (TAP) repair or valve-sparing (VS) approaches. Despite these advancements, ongoing debates persist regarding the optimal method of RVOT reconstruction, particularly in tailoring approaches to the severity of the disease in individual patients. [5–7]

The pulmonary valve annulus (PVA) z-score is widely utilized as a key predictor for determining the need for TAP repair, although cutoff values vary globally due to differences in surgical preferences and institutional practices. When the PVA z-score is critically low, TAP repair is typically employed. However, a significant drawback of TAP is the disruption of pulmonary valve integrity, leading to progressive pulmonary regurgitation (PR). Minimizing the risk of postoperative PR during initial repair is essential, as a substantial proportion of repaired ToF patients may require late pulmonary valve replacement to address complications such as right ventricular dilatation, ventricular arrhythmias, and sudden death. [5,8–10]

To mitigate the long-term sequelae of PR and the need for reoperations, contemporary surgical approaches emphasize pulmonary valve preservation or valve-sparing (VS) techniques as a less invasive approach during primary ToF repair. Although VS strategies have evolved significantly, they are associated with a trade-off: several studies show higher risk of residual pulmonary stenosis (PS) in the early postoperative but a reduced incidence of PR. [9–12]

This meta-analysis aims to consolidate institutional experiences worldwide into a unified study, enabling the development of a more accurate preoperative cut-off value for the PVA z-score as a less invasive approach. By synthesizing data from various centers, this approach seeks to establish a standardized predictive threshold that can guide surgical decision-making. The goal is to minimize complications associated with different operative techniques by leveraging the collective expertise and outcomes reported across institutions over recent years.

## 2. Materials and Methods

A systematic review and meta-analysis was rigorously undertaken to precisely evaluate the role of the PVA z-score within the context of preoperative assessment for surgical intervention, specifically focusing on valve-sparing procedures and techniques employing transannular patch reconstruction. The comprehensive literature search was executed over a defined period, commencing in February 2025 and concluding in October 2025, with the final update and data extraction performed on October 31, 2025. This entire process strictly adhered to the methodological standards and reporting architecture stipulated by the Preferred Reporting Items for Systematic Reviews and Meta-Analyses (PRISMA) statement.

### 2.1. Eligibility Criteria

Studies were included in this systematic review if they met the following inclusion criteria:

1. Original research comparing VS and TAP techniques on RVOT ToF reconstruction including: randomized controlled trials or observational studies (retrospective and prospective cohort studies).
2. Relevance to the clinical PICO framework:

- Patients: Pediatric patients (age < 18 years old) diagnosed with Tetralogy of Fallot, eligible for primary corrective treatment.
- Intervention: Assessment of preoperative Z-scores in patients undergoing valve/annular-sparing (VS) RVOT reconstruction. VS procedure defined as any surgical reconstruction of the RVOT where the native pulmonary valve annulus is preserved (left intact). This category may includes techniques such as pulmonary valvotomy, commissurotomy, leaflet delamination, and infundibular patching (transatrial or transventricular) that terminates proximal to the pulmonary annulus.
- Comparison: Preoperative Z-scores in patients undergoing transannular patch (TAP) RVOT reconstruction. TAP procedure included any surgical reconstruction involving an incision across the pulmonary valve annulus to relieve stenosis, typically augmented with a patch. Techniques involving annular division and patch, even if accompanied by leaflet preservation strategies, were classified as TAP for the purpose of this analysis, as they involve the disruption of the native annular valve geometry.
- Outcome: Evaluation of the preoperative mean difference, grand means, and the cut-off value of the pulmonary valve annulus z-score for each surgical approach.

Meanwhile, several exclusion criteria were as follows:

1. Uncontrolled studies, such as descriptive studies, case series or case reports.
2. Publications not categorized as original research, including review articles or commentaries.
3. Studies with full-text articles not available in English, resulting in a language barrier.

### 2.2. Search and Selection for Studies

A systematic literature search was conducted across multiple databases, including PubMed, ScienceDirect, SpringerLink, Scopus, Central/Cochrane, and Google Scholar. Several records also were identified from other methods such as, websites and citations searching. The search strategy employed the following terms: (“tetralogy of Fallot” OR “ToF”) AND (“valve sparing” OR “valve-sparing” OR “annulus sparing”) OR (“trans annular patch” OR “transannular patch” OR “TAP”) AND “z-score”. Duplicate entries were excluded, and the remaining articles were independently screened for relevance based on their abstracts by six authors (DF, JS, SP, PB, BR, PU). Fulltext reviews of the shortlisted articles were then conducted by the other five authors (LH, MD, DP, NV), applying the predefined inclusion and exclusion criteria.

### 2.3. Extraction and Quality Assesment of Data

Data extraction was independently carried out by three authors (JS, SP, DF) using standardized forms. The extracted information included the author, year of publication, study design, sample size, participant age, weight, and PVA z-score. The primary variable analyzed in the meta-analysis was the preoperative PVA z-score, reported as the mean ± standard deviation (SD), for both patient groups undergoing VS and TAP RVOT reconstruction. The PVA z-score is defined as a standardized metric quantifying the deviation of the pulmonary annulus diameter from the mean of a healthy population, normalized for body surface area, weight, and age. [12]

The quality assessment of the included observational cohort studies was independently conducted by three authors (MD, DP, NV) using the Newcastle-Ottawa Scale (NOS). This tool evaluates studies based on three broad perspectives: the selection of the study groups, the comparability of the groups, and the ascertainment of the outcome of interest. Studies were awarded stars to assess quality, with a maximum possible score of 9. The results of the quality appraisal were summarized in tables, classifying studies as having good, fair, or poor quality based on their total scores. Publication bias was also assessed visually by inspecting the funnel plot for asymmetry. To statistically quantify this assessment, Egger’s linear regression test was performed. A p-value of < 0.05 was considered indicative of significant publication bias.

### 2.4. Statistical Analysis

The statistical analysis for this study was conducted using meta-analytic methods implemented in Review Manager (RevMan) version 5.4.1. The effect estimate was expressed as the mean difference (MD) in forest plot with its standard deviation (SD) and 95% confidence interval (CI). Heterogeneity among studies in meta-analysis was evaluated using the I² statistic, with threshold of >50% indicating significant heterogeneity, requiring a random-effect model. A fixed-effect model was employed when heterogeneity was low. Continuous variables were analyzed using the inverse-variance method. Statistical significance was determined using a two-tailed p-value, with a threshold of ≤ 0.05.

When data were reported as medians and interquartile ranges (IQRs), they were converted to means and standard deviations using the formula proposed by Wan et al. [13] These mean values from the meta-analysis were subsequently used to construct receiver operating characteristic (ROC) curves in SPSS version 27. The area under the curve (AUC) was calculated to assess the discriminatory power of significant parameters, and optimal cut-off values were determined based on the sensitivity and specificity at various thresholds. [14]

## 3. Results

### 3.1. Literature Search and Selection

A systematic search of electronic databases and supplementary sources yielded a total of 766 records. Following the removal of 325 duplicates, 429 records remained for title and abstract screening via databases. This initial screening led to the exclusion of 318 records. We subsequently attempted to retrieve the full text of the remaining 123 reports from databases and other methods; 48 were not available, leaving 75 articles for full eligibility assessment. During this phase, 41 studies were excluded, primarily because they failed to present preoperative z-score values in patients undergoing RVOT reconstruction or lacked a direct comparison between VS and TAP group. Ultimately, 34 studies were incorporated into the systematic review, 29 of which were included in the meta-analysis. The detailed process of the literature search and study selection is illustrated in **Figure 1**.

**Figure 1.**
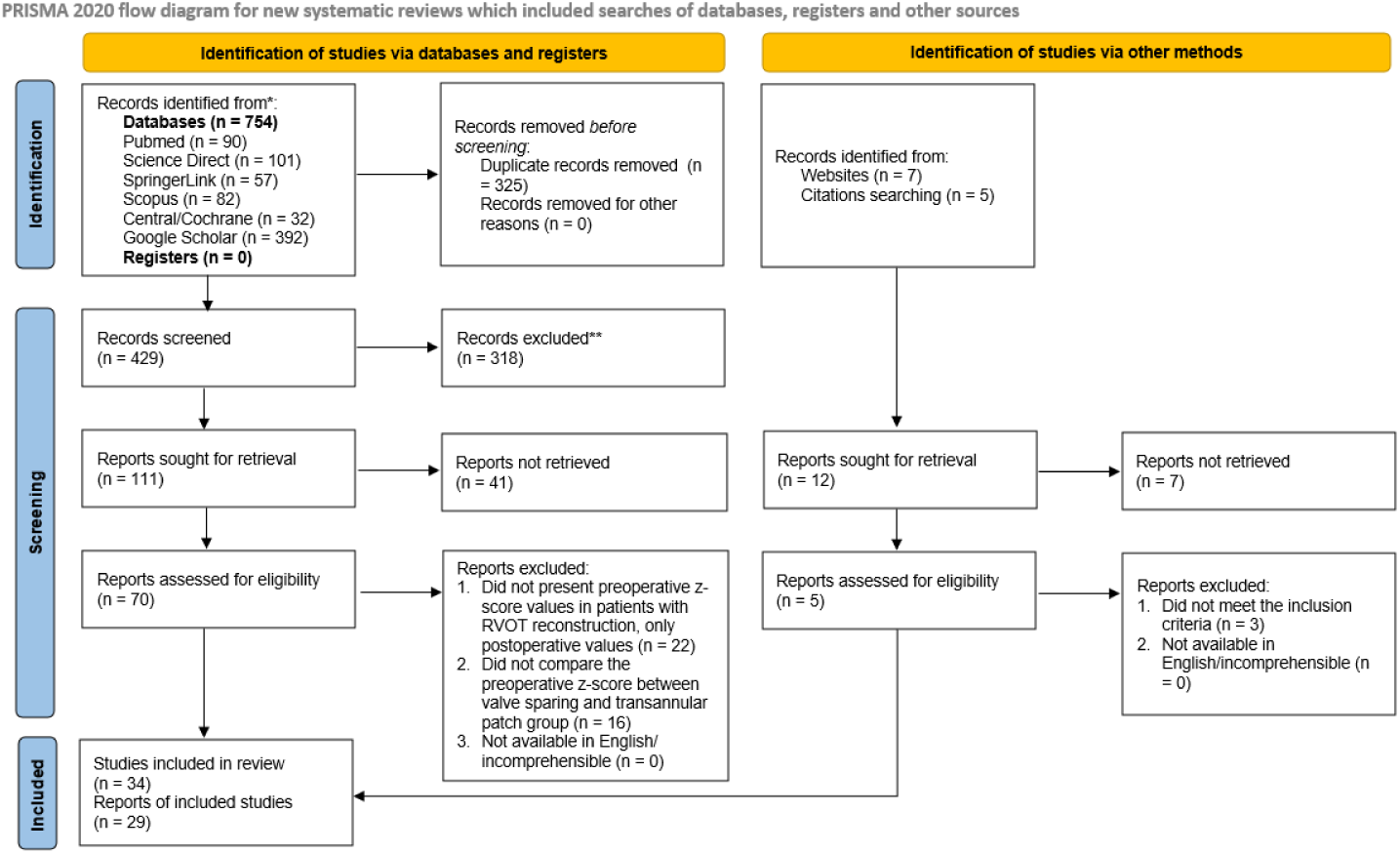
PRISMA 2020 flow diagram detailing the literature search and selection strategy.

### 3.2. Study Quality Assessment

The methodological quality of the 34 included studies was assessed using the Newcastle-Ottawa Scale (NOS) for cohort studies. Total quality scores ranged from 5 to 9 stars. Based on the established criteria, the majority of the studies (n = 22) were classified as good quality with a low risk of bias (scores of 7-9). The remaining 12 studies were categorized as fair quality with a moderate risk of bias, scoring between 5 and 6 stars. No studies were identified as having a high risk of bias (poor quality). Summary table detailing the assessment is presented in **Table A1.**

### 3.3. Characteristics of Studies

A total of 34 studies meeting the inclusion criteria were identified, then 29 of which included in numerical analysis comprising data from 5,806 patients with ToF. [3,5,7–12,15–40] Among these, 3,392 patients underwent RVOT reconstruction with VS, while 2,414 underwent TAP. Regarding study design, 31 studies (91.2%) were retrospective cohort studies, and 3 studies (8.8%) were prospective cohort studies. This systematic review encompasses data from three continents, with 17 studies originating from Asia (50%), 10 from North America (29.4%), and 7 from Europe (20.6%). Detailed study characteristics are presented in **Table 1**.

**Table 1.**
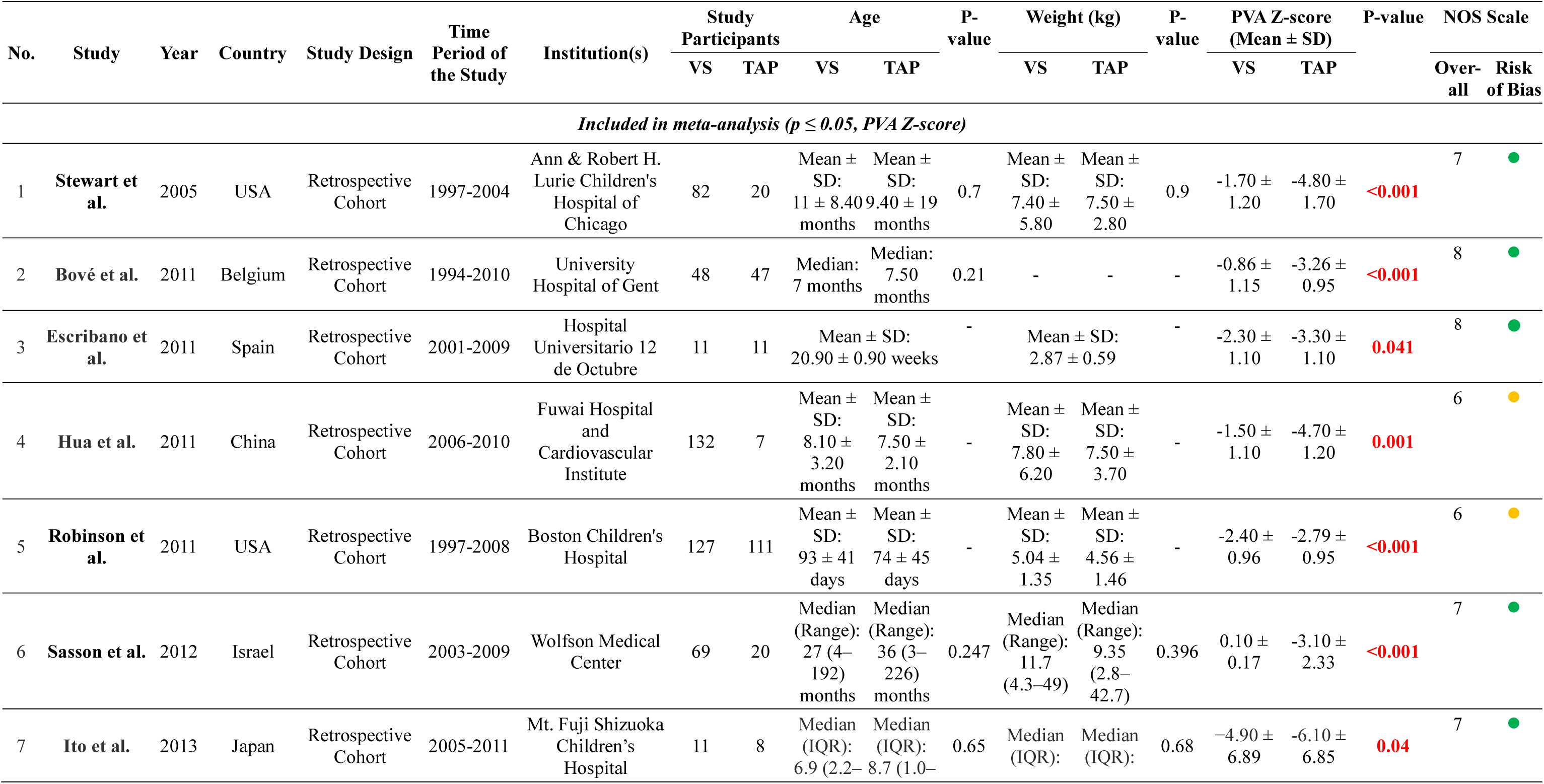
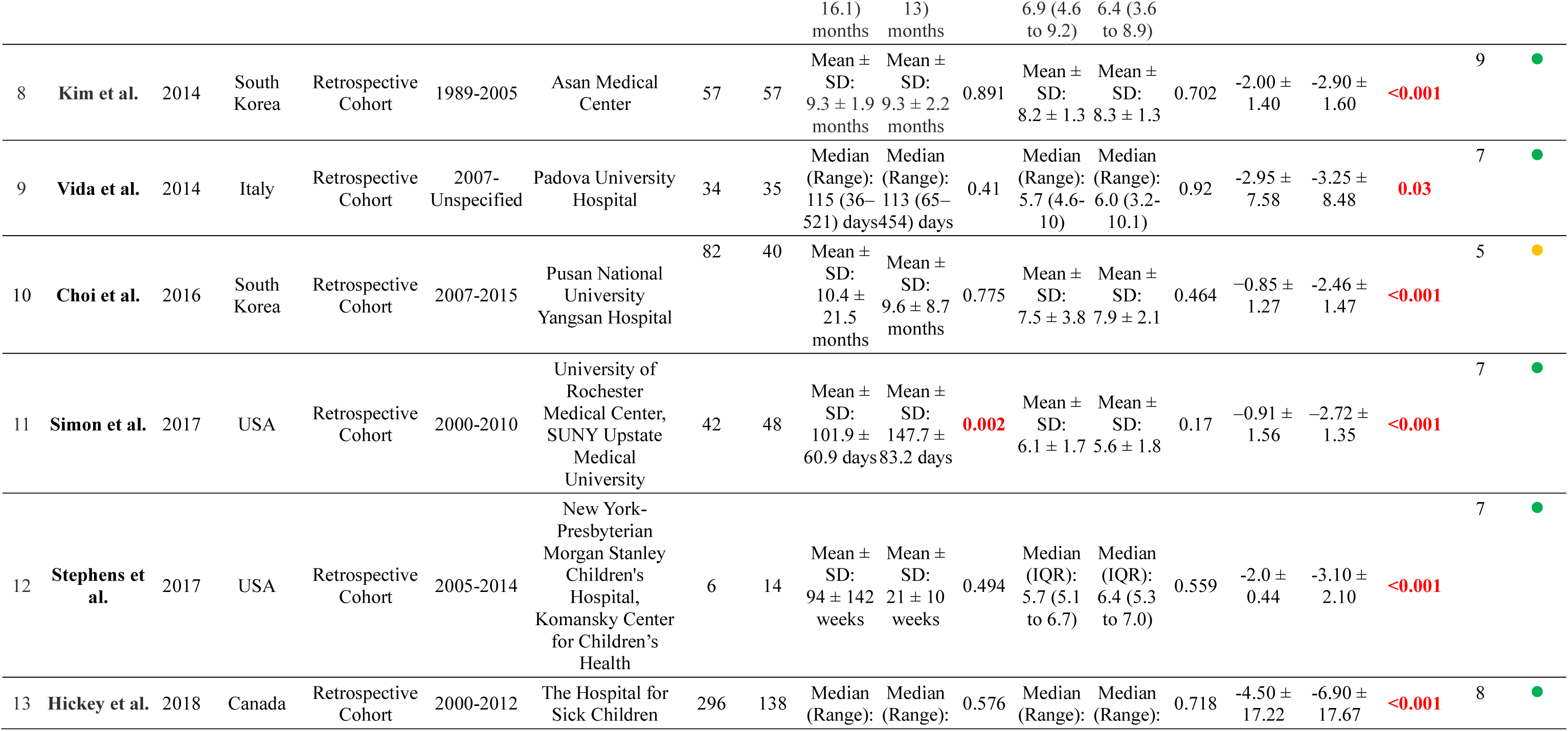
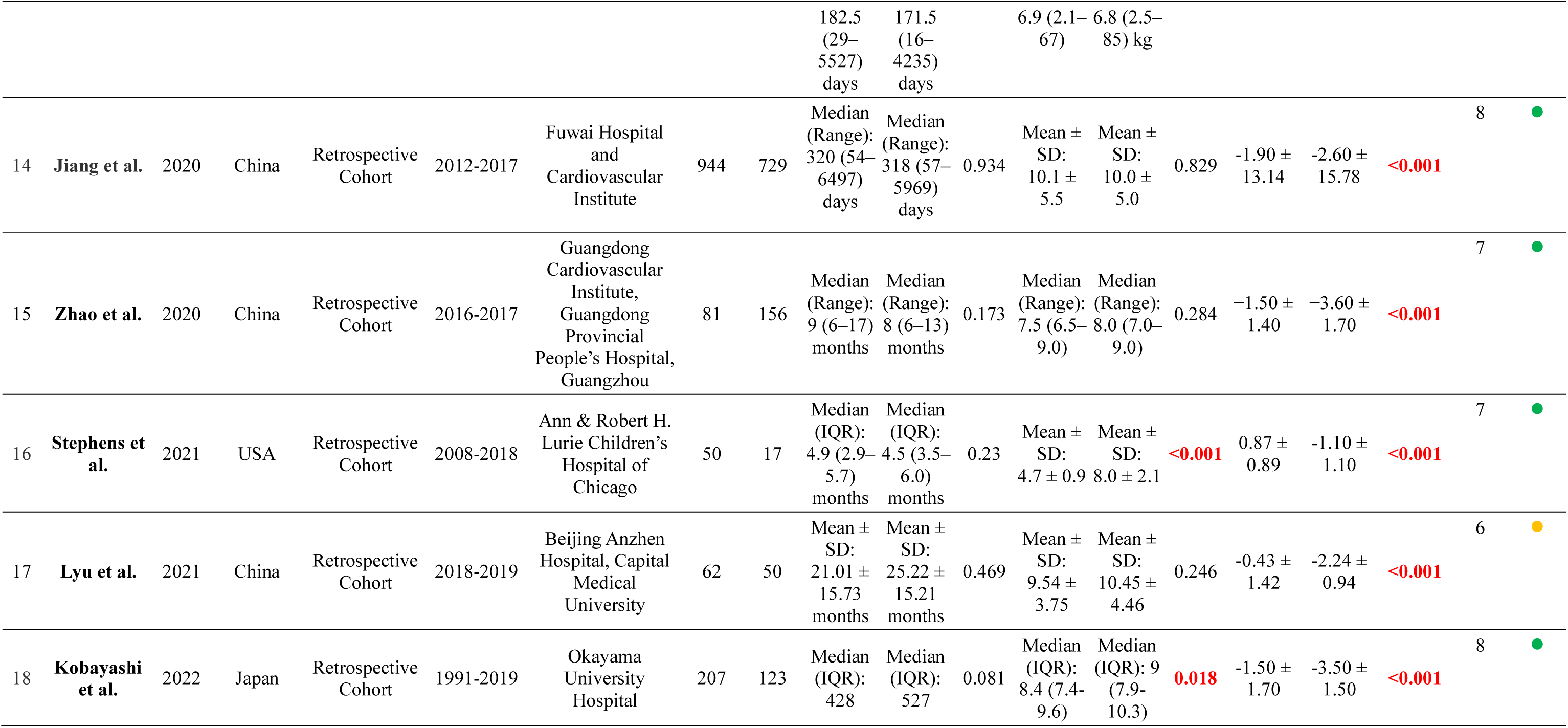
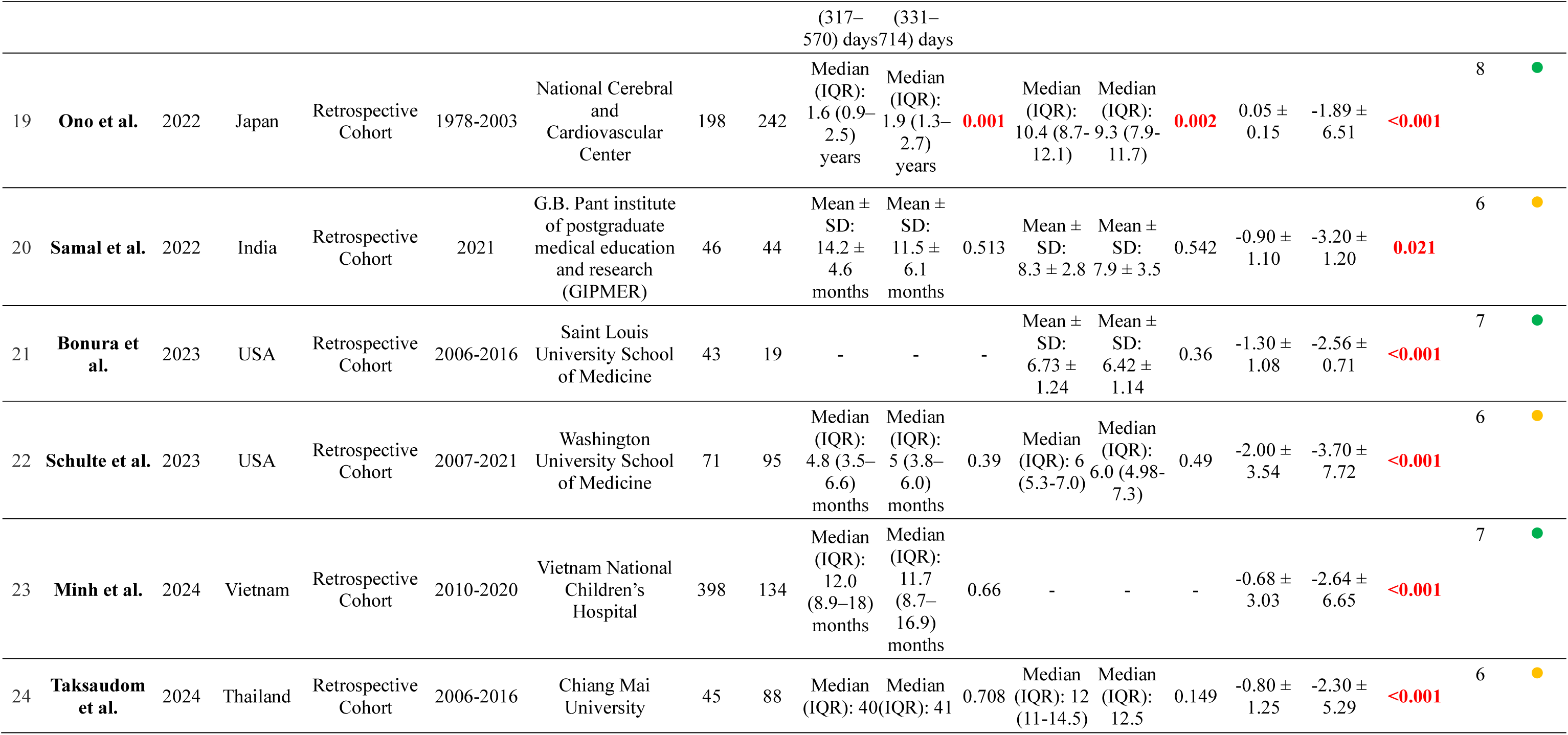
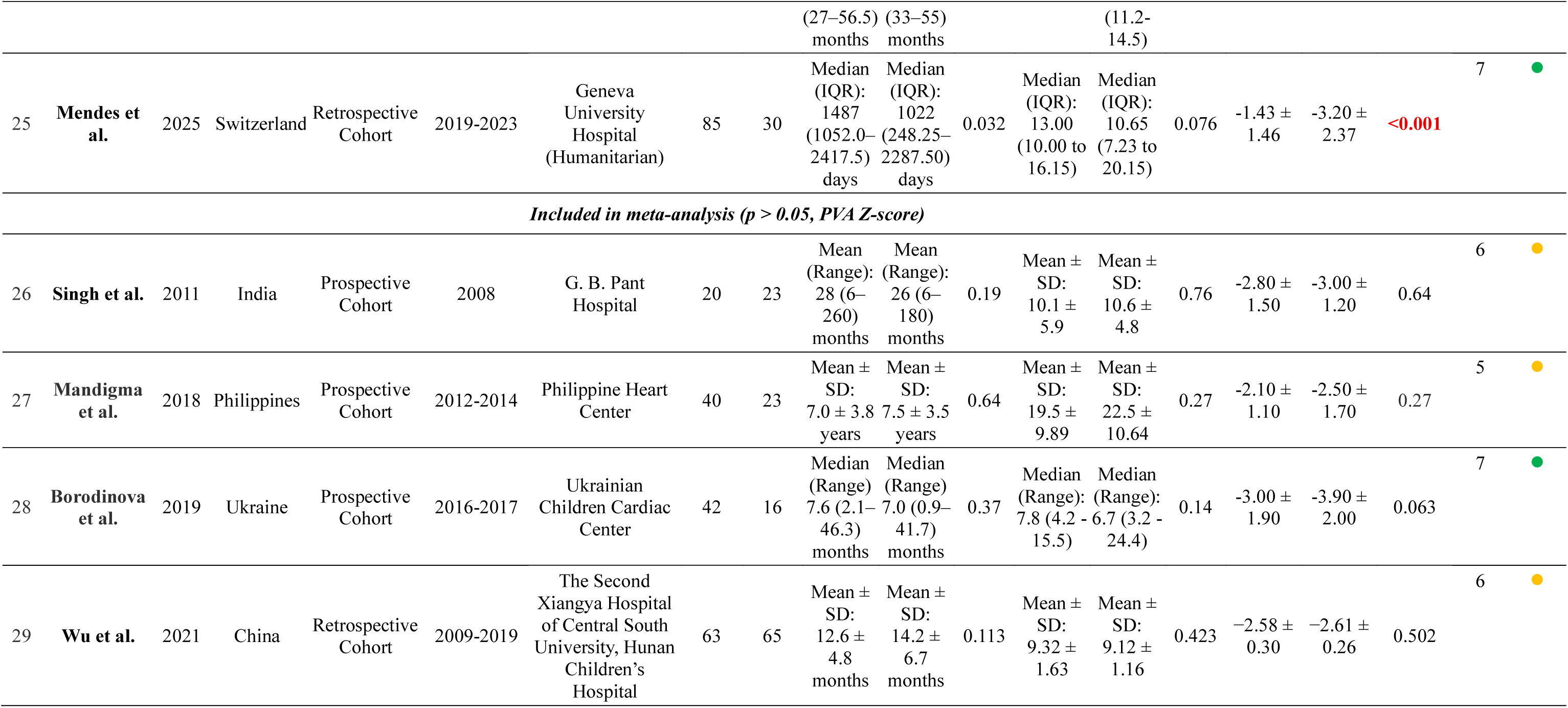
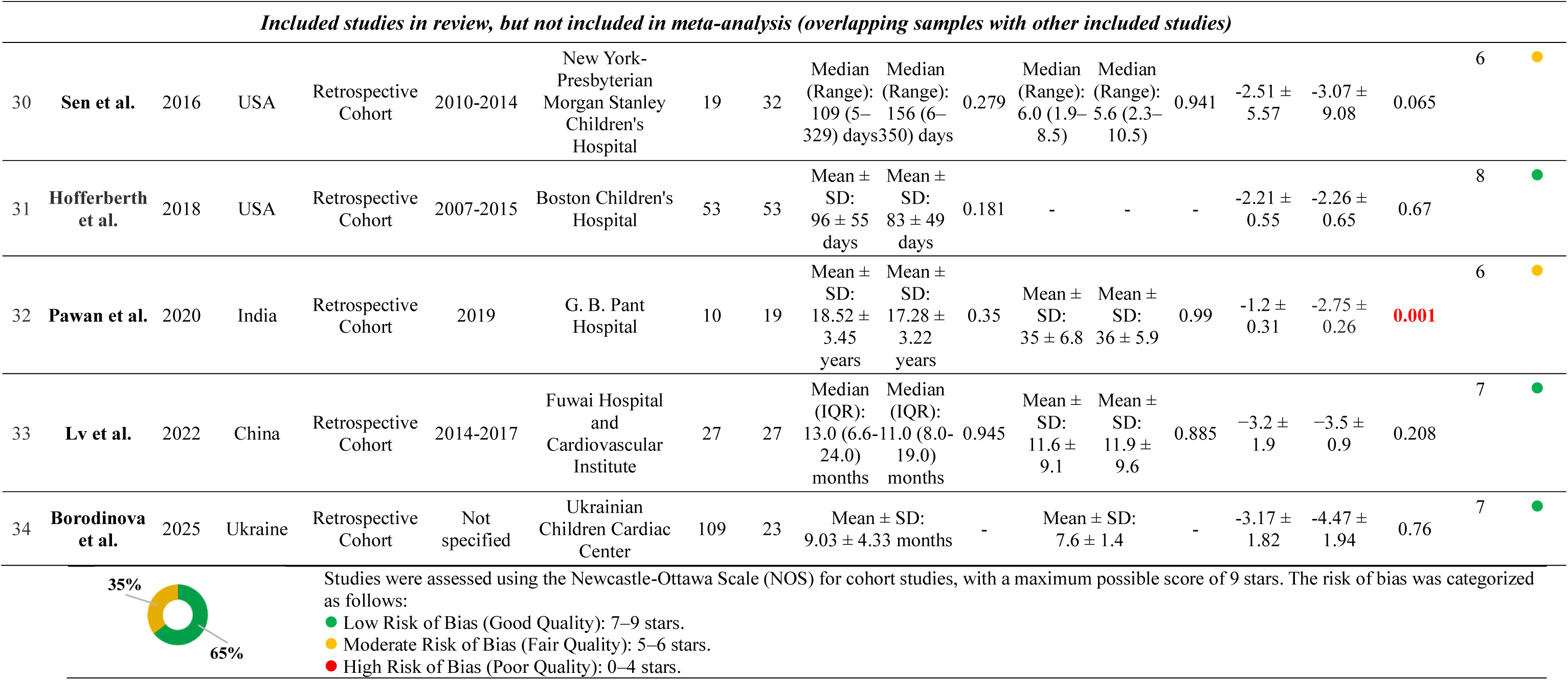
Characteristics of included studies.

### 3.4. Meta-Analysis Outcomes

#### 3.4.1. Standardized Mean Difference of Z-Score between VS and TAP Group

The quantitative analysis of the preoperative standardized mean difference (SMD) Z-score was summarized in **Figure 2**, with 29 inclusion studies. These studies encompassed 5,510 patients with significant p-values across 25 studies and 296 patients from 4 studies with nonsignificant p-values. The overall findings of the meta-analysis demonstrated a positive SMD, with a statistically significant p-value (p < 0.05), indicating an increase in preoperative Z-scores associated with the use of VS RVOT reconstruction (SMD: 1.01; 95% CI: 0.75 - 1.28; p < 0.001; I² = 95%). The high heterogeneity of the meta-analysis (I² > 50%) necessitated the use of a random-effects model. In the subgroup of studies with significant p-values, the analysis also revealed a positive mean difference, a significant overall p-value, and substantial heterogeneity (SMD: 1.14; 95% CI: 0.85–1.43; p < 0.001; I² = 95%).

**Figure 2.**
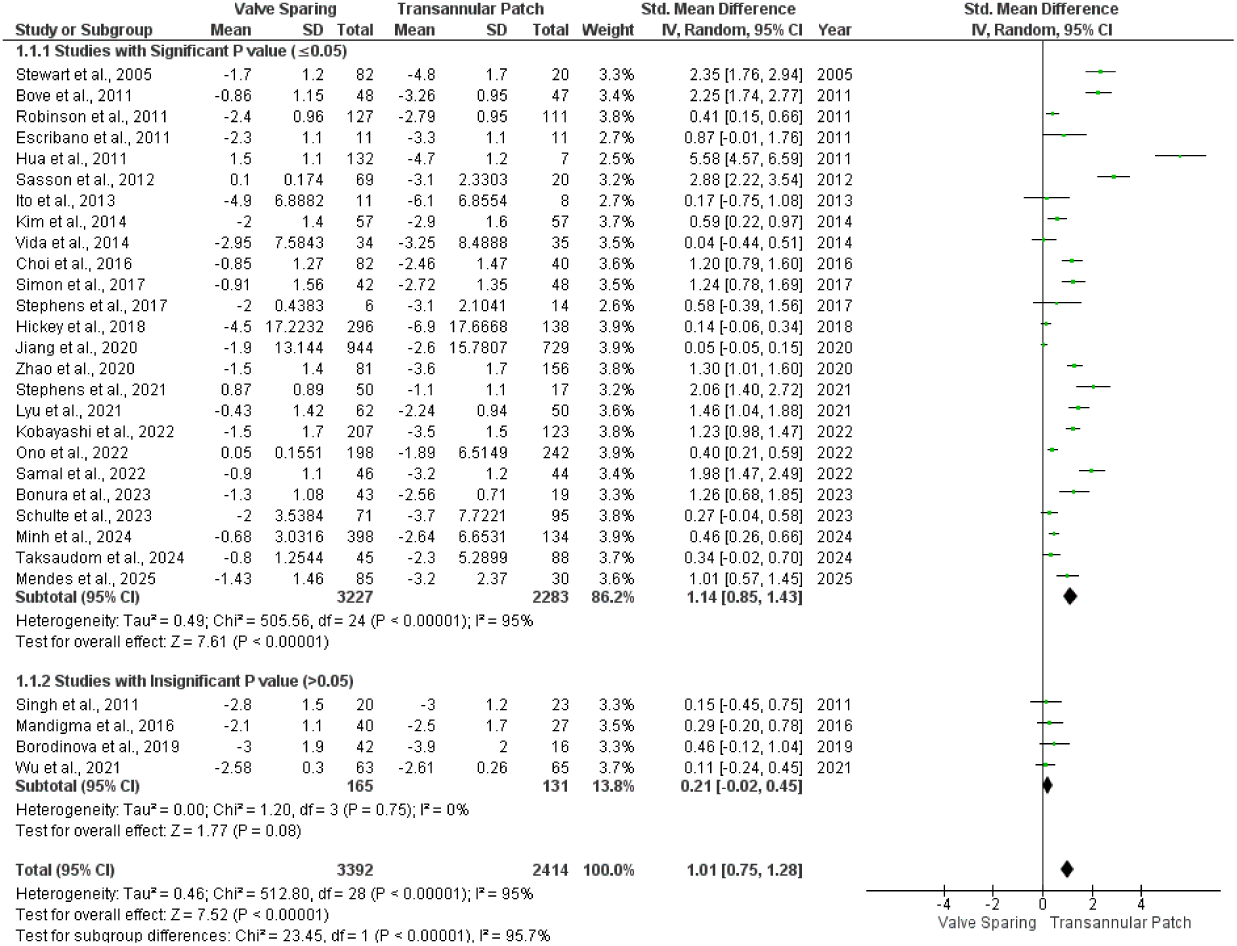
Forest plot illustrating the meta-analysis of preoperative Z-score mean differences stratified by the type of RVOT reconstruction.

#### 3.4.2. Preoperative Grand Mean of Z-Score in VS Group

This meta-analysis also evaluated the grand mean of the preoperative Z-score in the VS group, based on data from 29 studies, as illustrated in **Figure 3 (a)**. The grand mean preoperative Z-score, calculated from 3,392 patients in the VS group, was −1.39 (95% CI: −1.79 to −0.98; p < 0.001; I² = 100%). These results are statistically significant, as indicated by a p-value < 0.05. Due to the heterogeneity of this analysis, a random-effects model was employed to calculate the grand mean.

**Figure 3.**
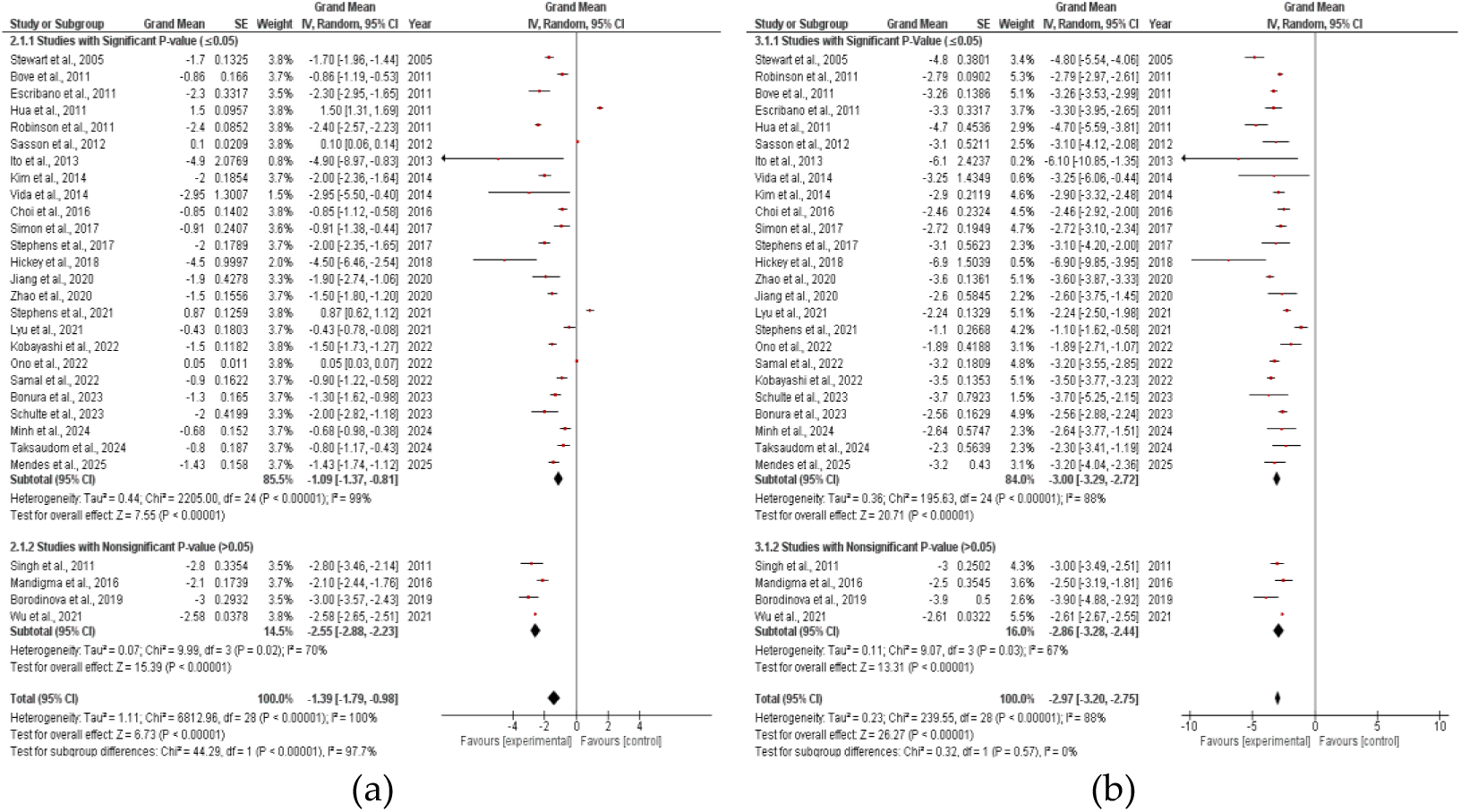
Forest plot graphics of preoperative grand mean Z-score in: (a) Patients undergoing valve-sparing technique; (b) Patients undergoing transannular patch technique.

#### 3.4.3. Preoperative Grand Mean of Z-Score in TAP Group

The grand mean of preoperative Z-scores in the TAP group, derived from 29 studies, is presented in **Figure 3 (b)**. Among 2,414 patients in this group, the grand mean preoperative Z-score was −2.97 (95% CI: −3.20 to −2.75; p < 0.001; I² = 88%). This result is statistically significant, with a p-value < 0.05. Due to the heterogeneity of the total analysis, instead of low heterogeneity in subgroup analysis (I² = 0%), a random-effects model was applied to calculate the grand mean in this meta-analysis.

#### 3.4.4. Preoperative Z-score Cut-Off Assessment for VS

An analysis was performed using the receiver operating characteristic (ROC) curve based on preoperative Z-score data from 29 included studies to determine the optimal Z-score cut-off for valve-sparing procedures, as shown in **Figure 4 (a)**. The ROC analysis demonstrated an area under the curve (AUC) value of 0.85 (>0.80; 95% CI, 0.84 – 0.86), indicating excellent discriminatory ability of the preoperative Z-score, with statistically significant results (p < 0.001).

**Figure 4.**
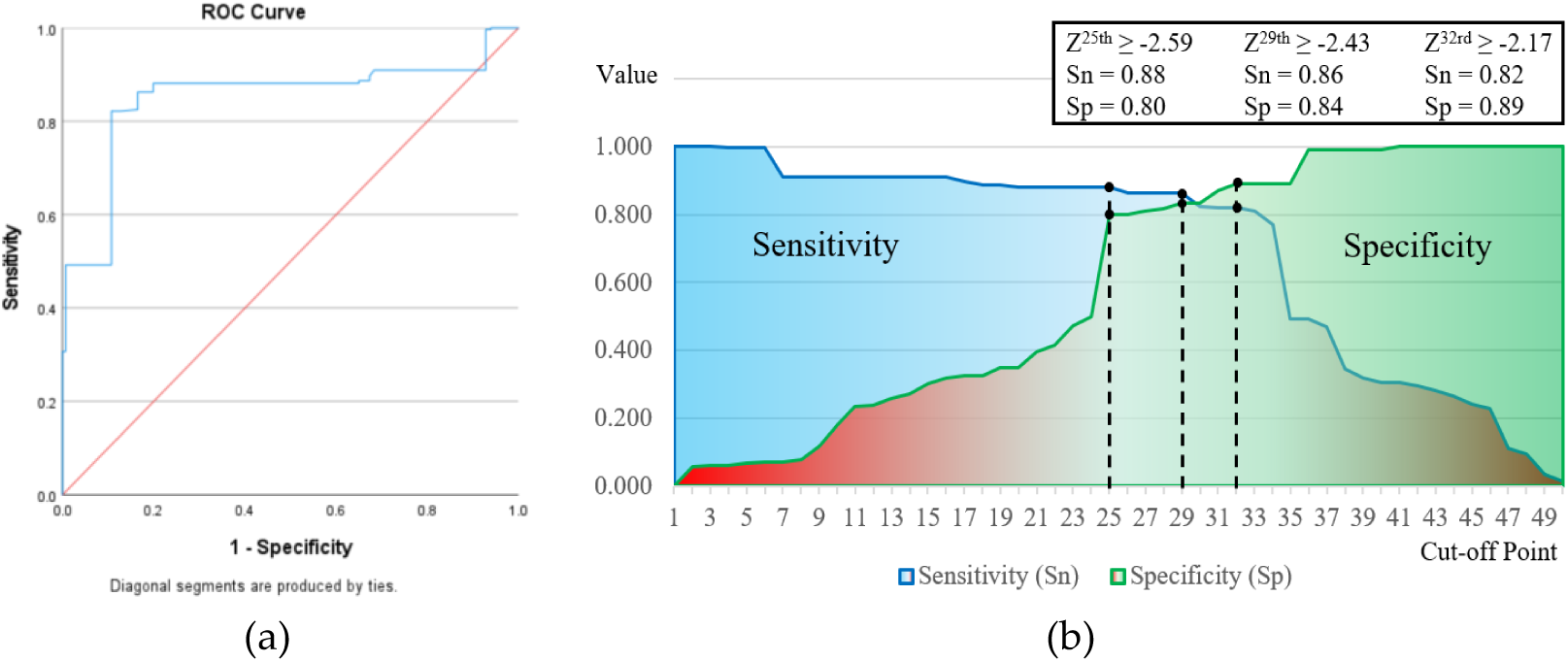
Preoperative Z-score cut off evaluation: (a) ROC curve analysis; (b) Sensitivity and specificity graphic for determining the optimal cut-off point.

The determination of the optimal Z-score cut-off values was based on identifying the best sensitivity and specificity at various points on the curve (> 80%). Three recommended Z-score cut-offs for guiding valve-sparing procedures were identified: Z-score ≥ −2.59 (sensitivity 88%, specificity 80%), Z-score ≥ −2.43 (sensitivity 86%, specificity 84%), and Z-score ≥ −2.17 (sensitivity 82%, specificity 89%). The detailed analysis of these cut-off points is presented in **Figure 4 (b)**.

## 4. Discussion

This systematic review and meta-analysis of 29 studies involving 5,806 patients demonstrates that the preoperative PVA z-score is a critical predictor for the feasibility of VS repair in ToF. Our analysis revealed a significantly higher preoperative PVA z-score in patients who underwent VS repair compared to those requiring a TAP (SMD: 1.01). Furthermore, we identified several suggestive diagnostic thresholds, for optimizing successful valve preservation. These findings provide a quantitative evidence base to guide surgical decision-making for VS techniques. The robustness of our data synthesis was confirmed by the absence of significant publication bias. Visual inspection of the funnel plot (Figure A1) showed a symmetrical distribution of studies, statistically supported by Egger’s test (p = 0.88). This indicates that the pooled results are not skewed by small-study effects. Therefore, recent surgical advancements have emphasized VS techniques aimed at preserving pulmonary valve function, minimizing pulmonary regurgitation (PR), and enhancing long-term survival and quality of life. [21,26,41]

The PVA size is a critical determinant in selecting the appropriate RVOT reconstruction strategy for patients with ToF. Patients with a markedly small PVA typically require TAP repair, whereas those with a sufficiently large PVA are more suitable candidates for VS approaches. However, for patients with intermediate PVA sizes, the choice of RVOT reconstruction method often depends on the surgeon’s expertise, preferences, or institutional practices, highlighting variability in clinical decision-making, without any standardized cut-off. [5,10,22]

This meta-analysis, coupled with ROC curve analysis of the preoperative PVA z-score, aims to provide insights into long-term outcomes, spanning over 20 years, across a large, global cohort of patients who have undergone either TAP or VS RVOT reconstruction strategies. This comprehensive approach allows for a more robust understanding of the long-term effects of these surgical methods. In this meta-analysis, the SMD in PVA Z-scores between patient groups undergoing RVOT reconstruction with VS and TAP was positive. This result was observed both across all studies and within studies with statistically significant p-values. These findings suggest that higher preoperative Z-score values are associated with a greater likelihood of employing VS techniques. [5,11,28,31]

This meta-analysis also included a grand mean z-score assessment across 29 studies comparing VS and TAP groups. The pooled grand mean PVA z-scores were −1.48 for the VS group and −2.93 for the TAP group. The higher grand mean z-score observed in the VS group aligns with the results of the mean difference analysis, supporting the association between larger PVA z-scores and the feasibility of VS procedures. [23,25,27]

While VS repair for ToF could help protect the right ventricle from volume overload, the potential for residual obstruction remains a concern. The application of PVA Z-score cut-off is remain inconsistent across countries, as reflected in the variability of reported median z-scores, with conflicting evidences in the literature.

Some studies suggest a PVA z-score of −2 as a threshold for TAP. [42] Meanwhile others, such as Stewart et al., found that a PVA z-score greater than −4 predicted successful pulmo-nary valve preservation with minimal recurrent obstruction. [15] Conversely, a Japa-nese study reported that even in patients with PVA z-scores below −4 (ranging from −6.3 to −4.3), valve-sparing surgery was successful in 58% of cases. [21] Furthermore, other studies indicate that valve-sparing techniques can be effectively performed in patients with PVA z-scores as low as −2.5, without increasing the risk for residual RVOT obstruction, compared to TAP. [3]

This study proposes three optimal cut-off values with the highest sensitivity and specificity for discrimination. The receiver operating characteristic (ROC) curve analysis demonstrated significant results, indicating an excellent discriminatory ability of the PVA z-score. The recommended cut-off values are as follows: z-score ≥ −2.59 (sensitivity 88%, specificity 80%), z-score ≥ −2.43 (sensitivity 86%, specificity 84%), and z-score ≥ −2.17 (sensitivity 82%, specificity 89%). Among these, the z-score ≥ −2.59 is preferred due to its higher sensitivity, enhancing the optimization of VS reconstruction. This recommendation aligns with recent studies emphasizing the benefits of maximizing VS techniques over TAP due to minimizing complications of TAP such as severe pulmonary insuficiency, cardiac arrhythmia, right ventricular dilatation, even sudden cardiac death. [9,21,22,36,37]

This meta-analysis has several limitations. First, the reliance on observational cohort studies introduces inherent selection bias, as surgical decisions were driven by intraoperative assessment rather than randomization. Second, high statistical heterogeneity (I2 > 50%) was observed, reflecting global variations in surgical techniques and patient demographics. Third, the pooled z-score results might be significantly influenced by methodological heterogeneity, as z-score values vary depending on the specific allometric method applied by each institution. Finally, while this study establishes the z-score as a feasibility predictor, it does not directly meta-analyze long-term functional outcomes such as right ventricular remodeling.

## 5. Conclusion

In conclusion, this meta-analysis demonstrates that VS repair is a viable alternative strongly associated with larger preoperative PVA z-scores. While TAP repair effectively relieves stenosis in Tetralogy of Fallot, it is associated with significant long-term pulmonary regurgitation. This meta-analysis identified an optimal z-score threshold of ≥ −2.59, offering reliable guidance for surgical decision-making with high diagnostic accuracy. Adopting this quantitative criterion allows for more precise surgical planning, promoting the broader use of valve-preservation strategies to minimize long-term complications and to improve patient survival.

## Author Contributions

Conceptualization, D.F., J.S., and S.P.; methodology, D.F. and J.S. ; software, J.S.; validation, D.F., S.P., P.B., and B.R.; formal analysis, J.S., N.V.; investigation, D.F., J.S., P.U., L.H.; resources, M.D., D.P.; data curation, J.S.; writing - original draft preparation, J.S.; writing - review and editing, D.F., J.S.; visualization, J.S., S.P.; supervision, P.B., B.R., M.D.; project administration, S.P., P.B.; funding acquisition, D.F. All authors have read and agreed to the published version of the manuscript.

## Supporting information

PRISMA Checklist

## Data Availability

All data produced in the present work are contained in the manuscript

## Abbreviations

The following abbreviations are used in this manuscript:

AUC: Area Under the Curve
CHD: Congenital Heart Disease
CI: Confidence Interval
IQR: Interquartile Range
MD: Mean Difference
NOS: Newcastle-Ottawa Scale
PICO: Patient, Intervention, Comparison, Outcome
PR: Pulmonary Regurgitation
PRISMA: Preferred Reporting Items for Systematic Reviews and Meta-Analyses
PS: Pulmonary Stenosis
PVA: Pulmonary Valve Annulus
RevMan: Review Manager
ROC: Receiver Operating Characteristic
RVOT: Right Ventricular Outflow Tract
SD: Standard Deviation
SMD: Standardized Mean Difference
SPSS: Statistical Package for the Social Sciences
TAP: Transannular Patch
ToF: Tetralogy of Fallot
VS: Valve-Sparing

## Appendix

**Table A1.** Quality assessment of included studies using Newcastle-Ottawa Scale.

| No. | Study and year | Selection |  |  |  | Comparability |  |  | Outcome |  | Overall | Risk of bias |
| --- | --- | --- | --- | --- | --- | --- | --- | --- | --- | --- | --- | --- |
|  |  | Representation of the exposed cohort | Selection of the non-exposed cohort | Ascertainment of exposure | Outcome not present at the start | Main factor | Additional factor | Assessment of outcome | Sufficiency of follow-up time | Adequacy of follow-up |  |  |
| 1 | Stewart *et al.*, 2005 | ★ | ★ | ★ | ★ | ★ | - | ★ | ★ | - | 7 | ● |
| 2 | Bové *et al.*, 2011 | ★ | ★ | ★ | ★ | ★ | - | ★ | ★ | ★ | 8 | ● |
| 3 | Escribano *et al.*, 2011 | ★ | ★ | ★ | ★ | ★ | - | ★ | ★ | ★ | 8 | ● |
| 4 | Hua *et al.*, 2011 | ★ | ★ | ★ | ★ | - | - | ★ | ★ | - | 6 | ● |
| 5 | Robinson *et al.*, 2011 | ★ | ★ | ★ | ★ | - | - | ★ | ★ | - | 6 | ● |
| 6 | Sasson *et al.*, 2012 | ★ | ★ | ★ | ★ | ★ | - | ★ | ★ | - | 7 | ● |
| 7 | Ito *et al.*, 2013 | ★ | ★ | ★ | ★ | - | - | ★ | ★ | ★ | 7 | ● |
| 8 | Kim *et al.*, 2014 | ★ | ★ | ★ | ★ | ★ | ★ | ★ | ★ | ★ | 9 | ● |
| 9 | Vida *et al.*, 2014 | ★ | ★ | ★ | ★ | - | - | ★ | ★ | ★ | 7 | ● |
| 10 | Choi *et al.*, 2016 | ★ | ★ | ★ | ★ | - | - | ★ | - | - | 5 | ● |
| 11 | Simon *et al.*, 2017 | ★ | ★ | ★ | ★ | - | - | ★ | ★ | ★ | 7 | ● |
| 12 | Stephens *et al.*, 2017 | ★ | ★ | ★ | ★ | - | - | ★ | ★ | ★ | 7 | ● |
| 13 | Hickey *et al.*, 2018 | ★ | ★ | ★ | ★ | ★ | - | ★ | ★ | ★ | 8 | ● |
| 14 | Jiang *et al.*, 2020 | ★ | ★ | ★ | ★ | ★ | ★ | ★ | ★ | - | 8 | ● |
| 15 | Zhao *et al.*, 2020 | ★ | ★ | ★ | ★ | - | - | ★ | ★ | ★ | 7 | ● |
| 16 | Stephens *et al.*, 2021 | ★ | ★ | ★ | ★ | ★ | - | ★ | ★ | - | 7 | ● |
| 17 | Lyu *et al.*, 2021 | ★ | ★ | ★ | ★ | - | - | ★ | - | ★ | 6 | ● |
| 18 | Kobayashi *et al.*, 2022 | ★ | ★ | ★ | ★ | ★ | - | ★ | ★ | ★ | 8 | ● |
| 19 | Ono *et al.*, 2022 | ★ | ★ | ★ | ★ | ★ | ★ | ★ | ★ | - | 8 | ● |
| 20 | Samal *et al.*, 2022 | ★ | ★ | ★ | ★ | - | - | ★ | - | ★ | 6 | ● |
| 21 | Bonura *et al.*, 2023 | ★ | ★ | ★ | ★ | ★ | - | ★ | ★ | - | 7 | ● |
| 22 | Schulte *et al.*, 2023 | ★ | ★ | ★ | ★ | - | - | ★ | ★ | - | 6 | ● |
| 23 | Minh *et al.*, 2024 | ★ | ★ | ★ | ★ | - | - | ★ | ★ | ★ | 7 | ● |
| 24 | Taksaudom *et al.*, 2024 | ★ | ★ | ★ | ★ | - | - | ★ | ★ | - | 6 | ● |
| 25 | Mendes *et al.*, 2025 | ★ | ★ | ★ | ★ | ★ | ★ | ★ | - | - | 7 | ● |
| 26 | Singh *et al.*, 2011 | ★ | ★ | ★ | ★ | - | - | ★ | - | ★ | 6 | ● |
| 27 | Mandigma *et al.*, 2016 | ★ | ★ | - | ★ | - | - | ★ | - | ★ | 5 | ● |
| 28 | Borodina *et al.*, 2019 | ★ | ★ | ★ | ★ | ★ | - | ★ | ★ | - | 7 | ● |
| 29 | Wu *et al.*, 2021 | ★ | ★ | ★ | ★ | - | - | ★ | ★ | - | 6 | ● |
| 30 | Sen *et al.*, 2016 | ★ | ★ | ★ | ★ | - | - | ★ | ★ | - | 6 | ● |
| 31 | Hofferberth *et al.*, 2017 | ★ | ★ | ★ | ★ | ★ | ★ | ★ | ★ | - | 8 | ● |
| 32 | Pawan *et al.*, 2020 | ★ | ★ | ★ | ★ | - | - | ★ | - | ★ | 6 | ● |
| 33 | Lv *et al.*, 2022 | ★ | ★ | ★ | ★ | ★ | - | ★ | ★ | - | 7 | ● |
| 34 | Borodina *et al.*, 2025 | ★ | ★ | ★ | ★ | - | - | ★ | ★ | ★ | 7 | ● |

Studies were assessed using the **Newcastle-Ottawa Scale (NOS)** for cohort studies, with a maximum possible score of 9 stars. The risk of bias was categorized as follows:
- **Low Risk of Bias (Good Quality):** 7–9 stars. - **Moderate Risk of Bias (Fair Quality):** 5–6 stars. - **High Risk of Bias (Poor Quality):** 0–4 stars.

**Figure A1.**
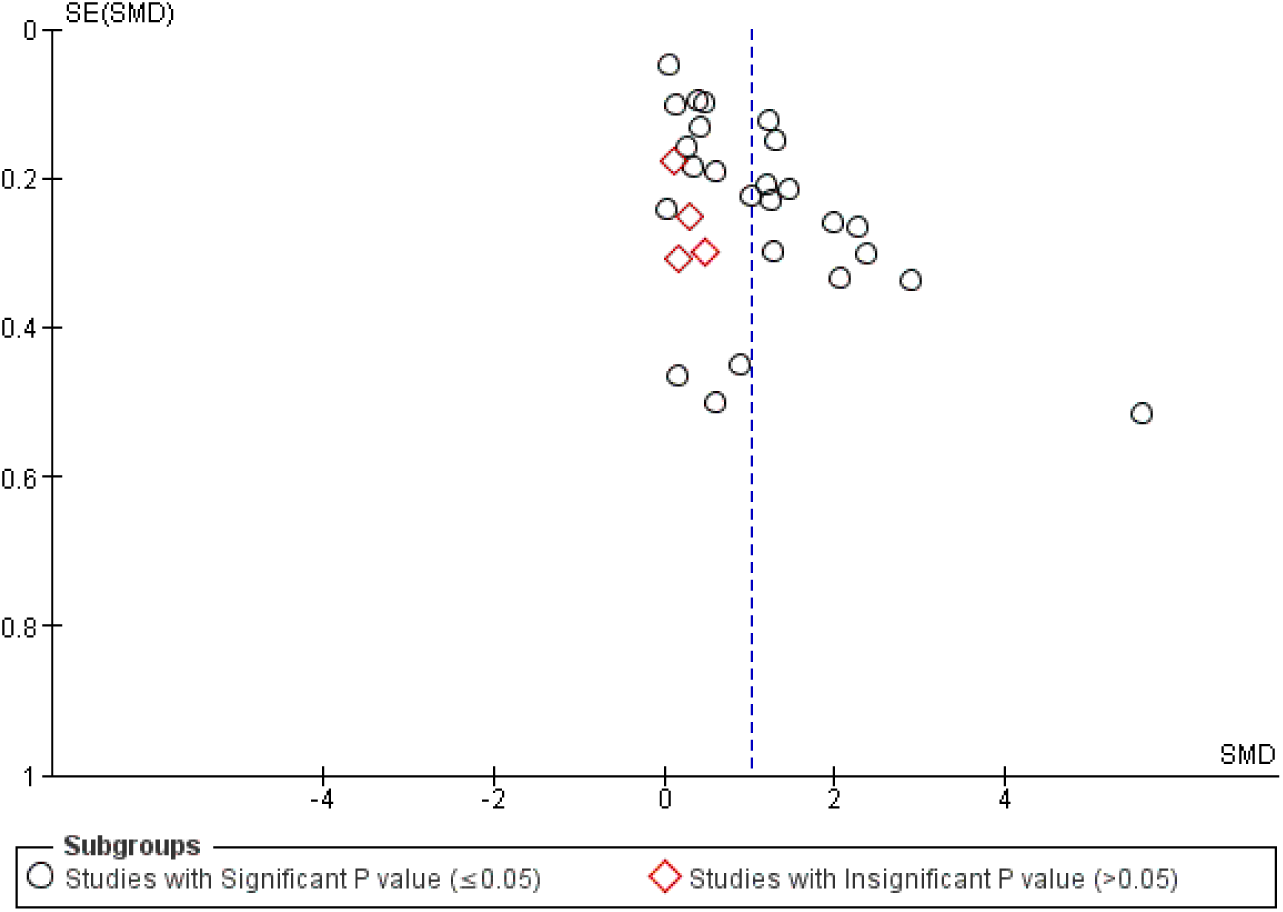
Funnel plot for preoperative PVA z-scores showing symmetrical distribution, confirming the absence of significant publication bias (Egger’s test p = 0.88).

